# Epidemiology and Determinants of Survival for Primary Intestinal Non-Hodgkin’s Lymphoma – A Population Based Study

**DOI:** 10.1101/2022.07.17.22277680

**Authors:** Vinit Singh, Dhairya Gor, Varsha Gupta, Aasems Jacob, Doantrang Du, Hussam Eltoukhy, Trishal Meghal

## Abstract

**Introduction:** Gastrointestinal tract is the most common site of extra nodal non-Hodgkin’s lymphoma (EN-NHL). Most of the published data have been on gastric NHL with limited studies on primary intestinal - Non-Hodgkin’s Lymphoma (PI-NHL) considering rare incidence. We performed epidemiological and survival analysis for PI-NHL from the Surveillance, Epidemiology, and End Results (SEER) 18 database.

**Methods:** A total of 9143 PI-NHL cases of age ≥18 years were identified from the SEER 18 database for the period 2000-2015. 8568 Patients were included for survival analysis. Cause specific Survival (CSS) and overall survival analysis (OS) were done for PINHL and PI-diffuse large B-cell lymphoma (PI-DLBCL) using gender, age of onset, treatment, histology, stage, and Year of diagnosis. Survival analysis was done by using cox-proportional hazard model and Kaplan Meier plot with log-rank test.

**Results:** The percentage of PI-NHL of all the intestinal cancers and extra nodal non-Hodgkin’s lymphoma were 1.35 %, and 10.52%, respectively. The age-adjusted incidence was 0.9145/100,000 population for the study population. PI-NHL was more common among patients aged≥60 Years, male and non-Hispanics whites. Majority of patients were diagnosed at stage 1 and 2 (74%), and DLBCL (44.8%) was the most common histology. In OS analysis, Significant increased risk of mortality was seen with T-cell NHLs vs. DLBCL (HR – 2.56), patients aged ≥60 vs <60 Years (HR – 2.87), stage 4 vs Stage 1 (HR – 1.93), male vs. female (HR- 1.17), with best outcome seen in patient treated with combination of chemotherapy and surgery vs. none (HR – 0.45). Similar results were seen in CSS and for primary intestinal DLBCL as well. Significant improvement in outcomes was observed for PI-DLBCL patients receiving chemotherapy with/without surgery.

**Conclusion:** Findings from our large, population-based study reveal PI-NHL is a rare type of intestinal malignancy with significant difference in survival based on histological and epidemiological characteristics.

## 1. Introduction

Non-Hodgkin’s lymphoma (NHL), a term encompassing various neoplasms of lymphoid origin, is the most common type of blood cancer in the USA(1). The most common site for extra-nodal Non-Hodgkin’s lymphoma (EN-NHL) is the gastrointestinal (GI) tract, constituting about 30-40% of the total NHL cases. Amongst primary GI tract lymphomas, gastric lymphoma is the most common site, followed by small intestinal lymphoma and colorectal lymphoma(2). Among types of non-Hodgkin’s lymphoma occurring in gastrointestinal tract, B cell lymphoma is more common than T cell lymphomas, with most T-cell lymphoma cases occurring in the ileocolic region of the intestine.

Primary intestinal non-Hodgkin’s lymphoma (PI-NHL) is a rare type of malignancy comprising about 2% of all gastrointestinal malignancies, and specifically about 2% of all small intestinal and 0.2% of all large intestinal malignancies(3,4). In a study done by the Danish lymphoma group, it was seen that over a period of 9 years between 1983 to 1991, the annual incidence rate for PINHL was 0.48 per 100,000, suggesting a very low incidence rate(5). However, there is a trend of gradual increase in the number of PI-NHL cases worldwide(2,6). Better diagnostic techniques, awareness about the disease, more colonoscopies and reporting have contributed to the increase in diagnoses of this disease. This has made it necessary to increase our focus on NHL with intestinal primary site. It should also be noted that for unknown reasons, up to 75% of primary GI tract lymphomas in the Mediterranean region and Middle East are PI-NHL and Burkitt lymphoma (BL), which has 50-fold higher incidence in Africa predominantly presents as obstructing lesion in the terminal ileum (7,8).

Over the years, several studies have been conducted to understand better epidemiological and survival parameters associated with primary gastrointestinal non-Hodgkin’s lymphoma (PGI-NHL). However, most of these studies primarily evaluated gastric NHL as opposed to PI-NHL, due to the lesser prevalence of the latter. Further, due to an overall lower incidence of affected PI-NHL patients, most of its observational and analytical studies have a relatively smaller sample size, making it challenging to derive conclusive evidence. In addition, a lower survival rate has been observed in intestinal lymphomas compared to gastric lymphoma (9), making it crucial to analyze the factors influencing survival rates in PI-NHL independently. Here, we present the epidemiological and survival data for PINHL from the Surveillance, Epidemiology, and End Results (SEER) 18 databases for 2000-2015.

## 2. Materials and Methods

This is a registry-based retrospective study executed by analysis of data from the SEER database for the years 2000 to 2015(10). The Population in SEER 18 represents 27.8% of the US population based on the 2010 US census. It includes cancer cases from 18 cancer registries across the United States including Alaska natives, Greater Georgia, Connecticut, Detroit (Metropolitan), Hawaii, Iowa, New Mexico, Rural Georgia, California excluding SF/SJM/LA, San Francisco-Oakland, San Jose-Monterey, Seattle, Utah, Kentucky, Los Angeles, Louisiana, New Jersey, and Atlanta (Metropolitan). The SEER database is a standard for population study in the United States with a case ascertainment rate of 98%. Individual patient-level data were extracted from the SEER 18 database using SEER*Stat software (version 8.0.5; Surveillance Research Program of the National Cancer Institute).

### 2.1. Study population

The database was queried for all patients diagnosed with PI-NHL between 2000 to 2015 based on EN-NHL ICD-O-3 recode and primary site code for small and large intestine (Primary site code C17.O – C21.8). Patients with age at diagnoses ≥ 18 were included in the study. Patients with unknown staging data, survival status and those diagnosed after autopsy were excluded from the study (Figure 1). Complete data collection process is described in detail in Supplement 1.

**Figure 1.**
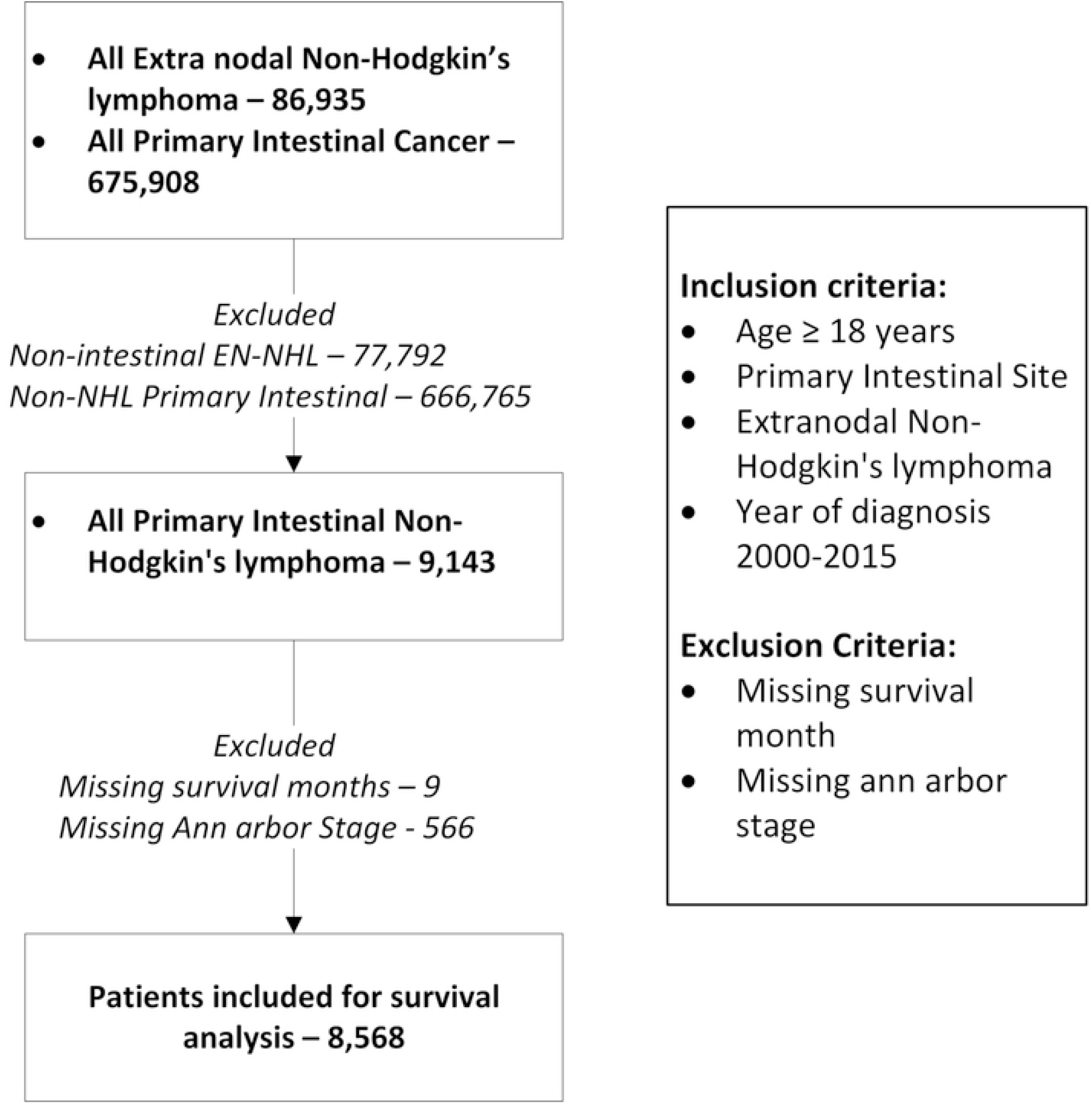
Description of patient selection and exclusion for the study. Patient listed in SEER 18 registries were included in the study.

The SEER registries collect data on patient characteristics, including age and year of diagnosis, origin, stage of diagnosis, gender, site, type of treatment, vital status, cause of death, and survival months at last follow-up or at death which were retrieved for the study. Associations between the demographic, clinical, and pathologic characteristics of patient with survival month were assessed. Age ≥ 60 is taken as a cutoff for analysis of the association between age and survival since it is considered an adverse prognostic factor as per the International Prognostic Index (IPI). Pathological information included stage at diagnosis and histological type of NHL. We also included year of diagnosis as one of the determinants in the survival analysis and was divided into three broad periods (2000-2005, 2006 -2010, and 2011-2015).

### 2.2. Statistical analysis

Descriptive statistics were used to describe patient baseline characteristics. Age-adjusted Incidence rates were calculated for study duration of 2000 – 2015 for age ≥ 18 years using SEER*Stat software. Age-adjusted incidence rate ratio were calculated for gender and ethnicity.

Overall survival (OS) and Cause-specific survival (CSS) were calculated as the time from diagnosis to death from any cause and from time of diagnosis to death from PI-NHL, respectively. Survival curves were plotted using Kaplan-Meier plots and survival analysis was performed using a log-rank test. Life tables were constructed to analyze the one-year, five-year and 10-year OS and CSS and were shown as percentages with a 95% confidence interval (95% CI). Patients were censored at the last follow-up in SEER, death or on December 31, 2018, whichever came first.

Hazard ratio was calculated by Multivariate cox-proportional hazard model. To identify the determinant of outcome, all the covariates were analyzed in a univariate cox - proportional hazard model for overall and cause specific survival. Those covariates which were significant for the univariate cause specific and overall cox-proportional hazard survival model were fitted into the multivariate model to analyze the effect of each covariate independent of the others. DLBCL was taken as reference for survival analyses considering most common type of NHL in the intestinal tract. We also calculated the survival factor for primary intestinal DLBCL considering it is the most common intestinal non-Hodgkin’s lymphoma in both small and large intestine.

. All the covariates were normalized for age, gender, and staging based on significance during univariate analysis. The strength of association between each predictor and survival was expressed as a hazard ratio (HR) along with a 95% CI. All tests were 2-sided, and a p-value of 0.05 was considered statistically significant. All the data analyses were done in Stata/IC 16.1 and R studio with R version 4.0.1 using “survival”, “surviminer” and ‘dplyr” packages.

## 3. Results

For the study period, 9143 patients age ≥ 18 years were identified as PI-NHL. 86,935 patients with EN-NHL, and 675,908 patients with any type of primary intestinal cancers were also identified. PI-NHL formed 1.35% (95% CI 1.32 – 1.38) of all intestinal cancers and 10.52% (95% CI 10.31 – 10.72) of all EN-NHL. The age-adjusted incidence of PI-NHL was 0.9145 per 100,000 individuals for age ≥ 18 for the study duration and were seen more in male and non-Hispanic white (NHW) population **(Table 1**). There was no significant change in the age adjusted incidence during the study duration. Among the 9143 patients with PI-NHL, 566 patients without Ann Arbor stage data and nine without survival data were excluded **(Supplement 1)**. We have summarized the demographic details in **Table 2**.

**Table 1:** Age adjusted incidence for PI-NHL for Year 2000-2015 for age ≥ 18. ¥ Arranged in descending order of Age-adjusted incidence rate

|  |  | Age Adjusted Incidence<br>(per 100,000<br>population) | Rate Ratio | P-Value |
| --- | --- | --- | --- | --- |
| **Study population**<br>**(Age ≥ 18, Year 2000-2015)** |  | 0.914 (0.896 – 0.934) | - | - |
| **Sex<sup>¥</sup>** |  |  |  |  |
|  | Male | 1.24 (1.207 – 1.274) | Reference | - |
|  | Female | 0.651 (0.629 -673) | 0.5246 (0.5026 – 0.5475) | 0.00 |
| **Ethnicity<sup>¥</sup>** |  |  |  |  |
|  | Non-Hispanic<br>white | 0.954 | Reference | - |
|  | Non-Hispanic<br>asia-pacific<br>islanders | 0.923 (0.859 – 0.991) | 0.9677 (0.8964 – 1.0433) | 0.40 |
|  | Hispanic | 0.873 (0.818 – 0.932) | 0.9156 (0.8533 – 0.9812) | 0.01 |
|  | Non-Hispanic<br>American<br>Indian | 0.662 (0.47 -0.904) | 0.6946 (0.4921 – 0.9491) | 0.02 |
|  | Non-Hispanic<br>black | 0.589 (0.540-641) | 0.6175 (0.5644 – 0.6743) | 0.00 |

**Table 2.** Demography, and clinical characteristics of PINHL patients in the study

| Characteristics |  | n (% or range) |
| --- | --- | --- |
| **Total** |  | 8568 |
| **Age** | Median (years) | 66.5 (55- 77) |
|  | <60 years | 2933 (34.23) |
|  | ≥60 years | 5635 (65.77) |
| **Gender** |  |  |
|  | Female | 3,352 (39.12) |
|  | Male | 5,216 (60.88) |
| **Ethnicity** |  |  |
|  | Non-Hispanic white | 6240 (72.83) |
|  | Non-Hispanic black | 533 (6.22) |
|  | Hispanic | 967 (11.29) |
|  | Others | 828 (9.66) |
| **Staging** |  |  |
|  | Stage 1 | 4,013 (46.84) |
|  | Stage 2 | 2,329 (27.18) |
|  | Stage 3 | 456 (5.32) |
|  | Stage 4 | 1,770 (20.66) |
| **Site** |  |  |
|  | Small Intestine | 4,696 (54.81) |
|  | Large Intestine | 3,872 (45.19) |
| **Histology** |  |  |
|  | Follicular Lymphoma | 1,563 (18.24) |
|  | MALTOMA | 1,160 (13.54) |
|  | DLBCL | 3835 (44.76) |
|  | Mantle cell Lymphoma | 447(5.22) |
|  | Burkitt Lymphoma | 344 (4.01) |
|  | Other B-Cell Lymphoma | 232 (2.71) |
|  | T-Cell Non-Hodgkin's Lymphoma | 392 (4.58) |
|  | Non-Hodgkin's Lymphoma, NOS | 595 (6.94) |
| **Treatment** |  |  |
|  | Chemotherapy only | 2018 (23.55) |
|  | Surgery only | 2446 (28.55) |
|  | Surgery and Chemotherapy | 2506 (29.25) |
|  | None | 1598 (18.65) |
| **Year of Diagnosis** |  |  |
|  | 2000 – 2005 | 2,911 (33.98) |
|  | 2006 - 2010 | 2,815 (32.85) |
|  | 2011 - 2015 | 2,842 (33.17) |

### 3.1. Patient characteristics

PI-NHL was more common among males compared to females (60.9% vs. 39.1%). Majority (65.8%) of the PI-NHL occurred in patients aged ≥60 years. Majority of the patients were non-Hispanic whites (72.83%), followed by Hispanics (11.29%) and African Americans (6.22%). More than seventy percent of PI-NHL were diagnosed at early stages (stage 1 and 2). The combination of surgery and chemotherapy (29.25%) was the most common approach used for the treatment. Only surgery or chemotherapy were used in 28.55% and 23.55 % of patients, respectively. No intervention was done in 18.65% of the patients.

### 3.2. Histological classification of PINHL

The histological distribution of PINHL is summarized in **Table 3**. The majority of the PINHL were B-cell lymphoma, with Diffuse large B-Cell lymphoma (DLBCL) (44.76%) being the most common histology followed by follicular lymphoma (18.24%) and MALToma (13.54%). T-cell lymphoma constituted 4.58% of the total cases and 77.8% cases involved small intestine. Majority (75.8%) of follicular lymphoma had small intestine as primary site while mantle cell lymphoma (81.7%) and MALTOMA (61.8%) were more common in large intestine.

**Table 3.** Distribution of histological subtypes of non-Hodgkin’s lymphoma across the intestinal tract

| **Histological subtype of NHL** | **Total** | **Small intestine** | **Large intestine** |
| --- | --- | --- | --- |
| Follicular Lymphoma | 1,563 (18.24) | 1,185 (25.23) | 378 (9.76) |
| MALTOMA | 1,160 (13.54) | 443 (9.43) | 717 (18.52) |
| DLBCL | 3835 (44.76) | 2,138 (45.53) | 1,697 (43.83) |
| Mantle cell Lymphoma | 447(5.22) | 82 (1.75) | 365 (9.43) |
| Burkitt Lymphoma | 344 (4.01) | 166 (3.53) | 178 (4.60) |
| Other B-Cell Lymphoma | 232 (2.71) | 80 (1.70) | 152 (3.93) |
| T-Cell Non-Hodgkin's Lymphoma | 392 (4.58) | 305 (6.49) | 87 (2.25) |
| Non-Hodgkin's Lymphoma, NOS | 595 (6.94) | 297 (6.32) | 298 (7.70) |

### 3.3. Survival analysis

OS and CSS were calculated. One-year, 5-year and 10-year survival probabilities are listed in **Table 4**. Median survival in the overall study population was 111 months (95% CI 105 -117), with a non-significant difference with HR 1.00 (95% CI 0.99 – 1.1, p-value 0.14) between small intestine (median survival: 117 months, 95% CI 109 – 128 months) and large intestinal lymphomas (median survival: 103 months, 95% CI 98 -112 months). Survival data are summarized in Table 5. Univariate Hazard ratio for all the covariates are summarized in the supplement 1, table 3. On the multivariate analysis, compared to patients aged < 60 years, patients aged ≥ 60 years had worse cause specific (HR 2.13, 95 CI 1.94 - 2.34) and overall survival (HR 2.87, 95% CI 2.65 – 3.10). Male gender conferred an increased risk of mortality for both CSS (HR 1.14, 95% CI 1.05-1.23) and OS (HR 1.17, 95% CI 1.10-1.24) compared to female gender. Stage 4 patients had a significantly higher risk of mortality compared to stage 1 disease (CSS HR 2.56, 95% CI 2.31-2.83; OS HR 1.93, 95% CI 1.79-2.09). Compared to DLBCL, Follicular Lymphoma (HR 0.29, 95% CI 0.26-0.32) and MALToma (HR 0.37, 95% CI 0.33 - 0.41) had better OS. T-Cell lymphoma however, had the worse OS outcomes compared to DLBCL (HR 2.56, 95% CI 2.28-2.88). BL conferred poor overall survival compared to DLBCL with hazard ratio trending towards statistical significance (HR 1.17, 95% CI 1.00-1.38).

**Table 4.** Overall survival and cause specific survival at 1 year, 5 year and 10 years for all PI-NHL patients.

|  | **Cause Specific Survival (95% CI)** | **Overall Survival (95% CI)** |
| --- | --- | --- |
| 1 Year Survival | 0.80 (0.796 – 0.813) | 0.76 (0.749 – 0.767) |
| 5 Year Survival | 0.71 (0.706 – 0.725) | 0.61 (0.600 – 0.621) |
| 10 Year Survival | 0.66 (0.654 – 0.677) | 0.48 (0.469 – 0.493) |

**Table 5.** Multivariate analysis of the survival factors for cause specific and overall survival for all PINHL patients.

|  | Variable | Cause Specific<br>Survival HR<br>(95% CI) | p-value | Overall Survival HR<br>(95% CI) | p-value |
| --- | --- | --- | --- | --- | --- |
| **Year of Diagnosis** |  |  |  |  |  |
|  | 2000-2005 | Reference |  | Reference |  |
|  | 2006-2010 | 0.87 (0.80 – 0.96) | 0.004 | 0.88 (0.82 – 0.95) | <0.001 |
|  | 2011-2015 | 0.72 (0.65 – 0.80) | <0.001 | 0.78 (0.72 – 0.84) | <0.001 |
| **Age** |  |  |  |  |  |
|  | <60 years | Reference |  | Reference |  |
|  | ≥60 years | 2.13 (1.94 – 2.34) | <0.001 | 2.87 (2.65 – 3.10) | <0.001 |
| **Gender** |  |  |  |  |  |
|  | Female | Reference |  | Reference |  |
|  | Male | 1.14 (1.05 – 1.23) | 0.006 | 1.17 (1.10 – 1.24) | <0.001 |
| **Stage** |  |  |  |  |  |
|  | Stage 1 | Reference |  | Reference |  |
|  | Stage 2 | 1.54 (1.40 – 1.71) | <0.001 | 1.26 (1.17 – 1.36) | <0.001 |
|  | Stage 3 | 1.88 (1.59 – 2.22) | <0.001 | 1.45 (1.27 – 1.66) | <0.001 |
|  | Stage 4 | 2.56 (2.31 – 2.83) | <0.001 | 1.93 (1.79 – 2.09) | <0.001 |
| **Histology** |  |  |  |  |  |
|  | DLBCL | Reference |  | Reference |  |
|  | Follicular Lymphoma | 0.19 (0.16 – 0.23) | <0.001 | 0.29 (0.26 – 0.32) | <0.001 |
|  | MALTOMA | 0.21 (0.17 – 0.25) | <0.001 | 0.37 (0.33 – 0.41) | <0.001 |
|  | Mantle Cell lymphoma | 0.60 (0.51 – 0.70) | <0.001 | 0.59 (0.52 – 0.67) | <0.001 |
|  | Other B-cell Lymphoma | 0.61 (0.49 – 0.77) | <0.001 | 0.68 (0.58 – 0.81) | <0.001 |
|  | NHL, NOS | 0.76 (0.66 – 0.89) | <0.001 | 0.83 (0.74 – 0.93) | 0.001 |
|  | Burkitt Lymphoma | 1.36 (1.13 – 1.64) | 0.001 | 1.17 (1.00 – 1.38) | 0.052 |
|  | T-Cell Non-Hodgkin's Lymphoma | 3.04 (2.67 – 3.46) | <0.001 | 2.56 (2.28 – 2.88) | <0.001 |
| **Treatment** |  |  |  |  |  |
|  | None | Reference |  | Reference |  |
|  | Surgery | 0.82 (0.73 – 0.93) | 0.008 | 0.96 (0.88 – 1.05) | 0.395 |
|  | Chemotherapy | 0.52 (0.46 – 0.60) | <0.001 | 0.59 (0.53 – 0.65) | <0.001 |
|  | Chemotherapy and surgery | 0.39 (0.34 – 0.44) | <0.001 | 0.45 (0.40 – 0.49) | <0.001 |

For treatment, there was no significant difference seen for OS between no treatment/observation and patients who only underwent surgery whereas CSS showed slight better outcome in patients with surgery with HR of 0.82 (95% CI 0.73 - 0.93). Patients who underwent chemotherapy and multimodality treatment with chemotherapy and surgery had better outcome than observation or surgery alone with minimal yet significant difference between themselves for both CSS and OS as shown in Table 5. Cause specific and overall survival of PI-NHL increased significantly over the years compared to 2000-2005 period. Overall survival and cause specific Kaplan-Meier curves were plotted for survival factors included in cox-proportional hazard model of PI-NHL and shown in **Figure 2 and Figure 3**, respectively. Survival analysis of PI-DLBCL showed similar trends as seen for broader PINHL and is summarized in **Table 6** with OS and CSS Kaplan Meier plots shown in **Figure 4 and 5** respectively.

**Figure 2.**
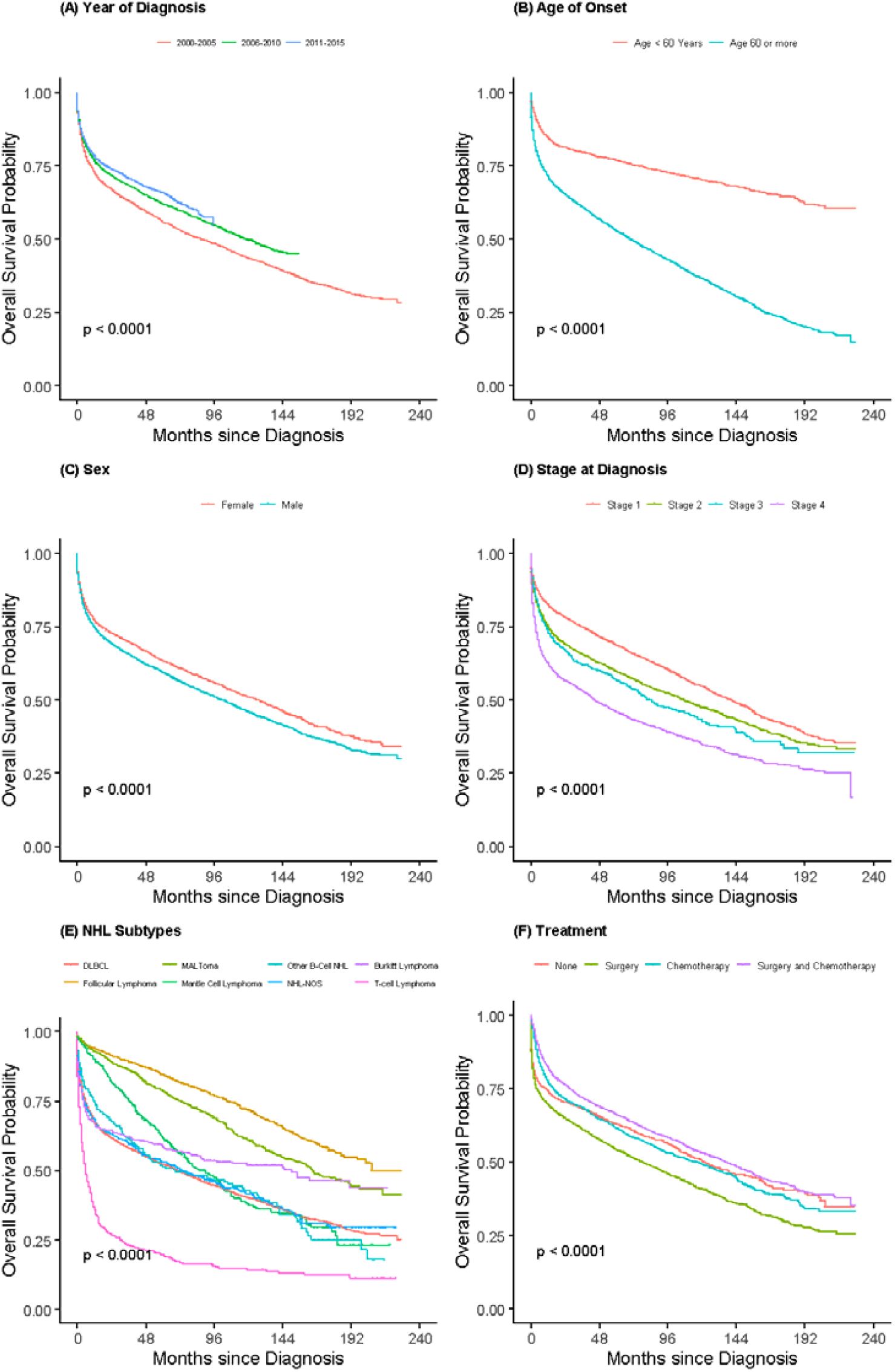
Kaplan Meier plots for Overall survival among all PI-NHL patients for effect (a) Year of Diagnosis and, (b) Age (c) Sex, (d) Stage of disease (e) NHL Subtypes, and (f) Treatment

**Figure 3.**
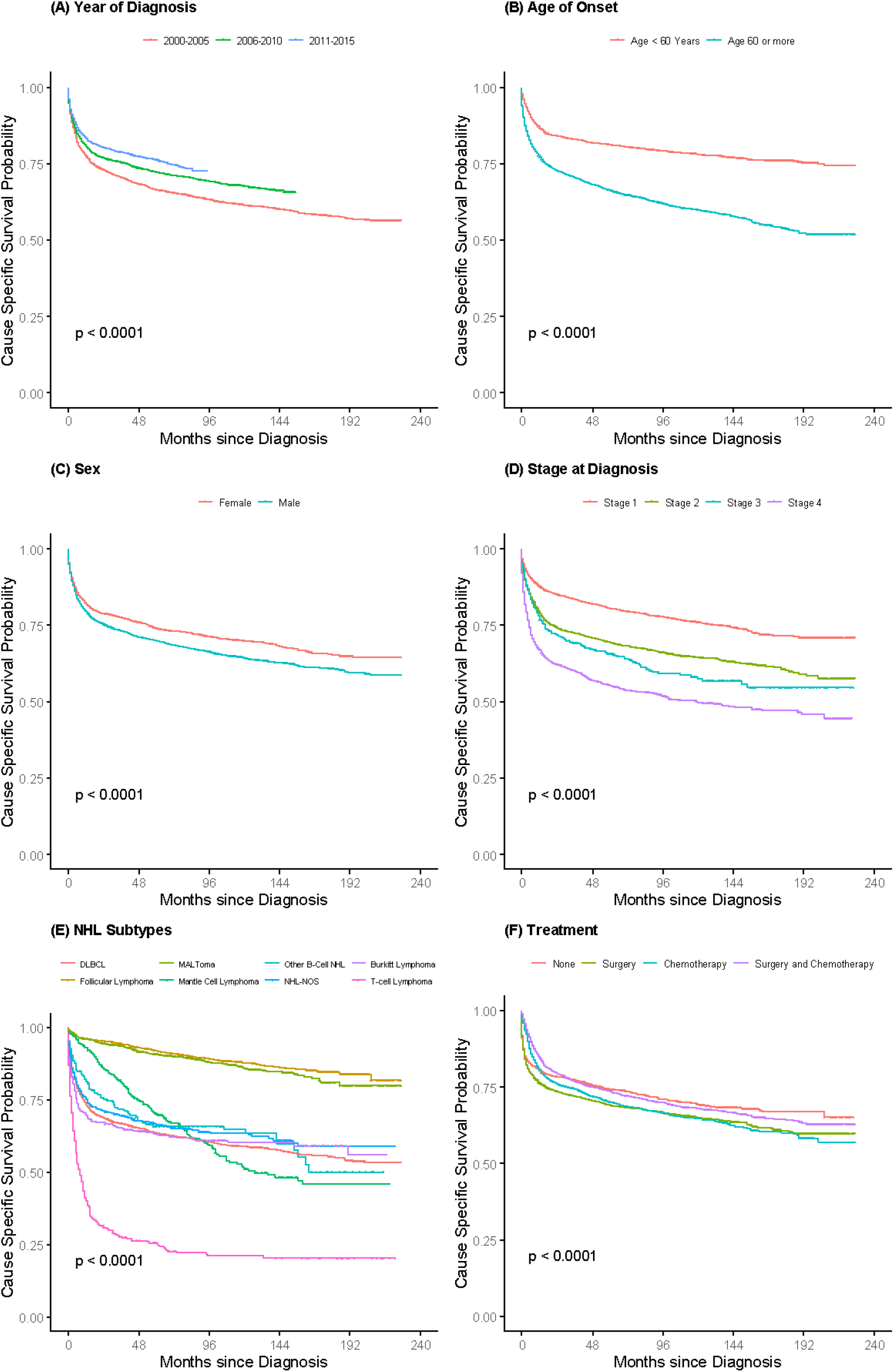
Kaplan Meier plots for Cause Specific survival among all PI-NHL patients for effect (a) Year of Diagnosis and, (b) Age, (c) Sex, (d) Stage of disease, (e) NHL Subtypes, and (f) Treatment.

**Figure 4.**
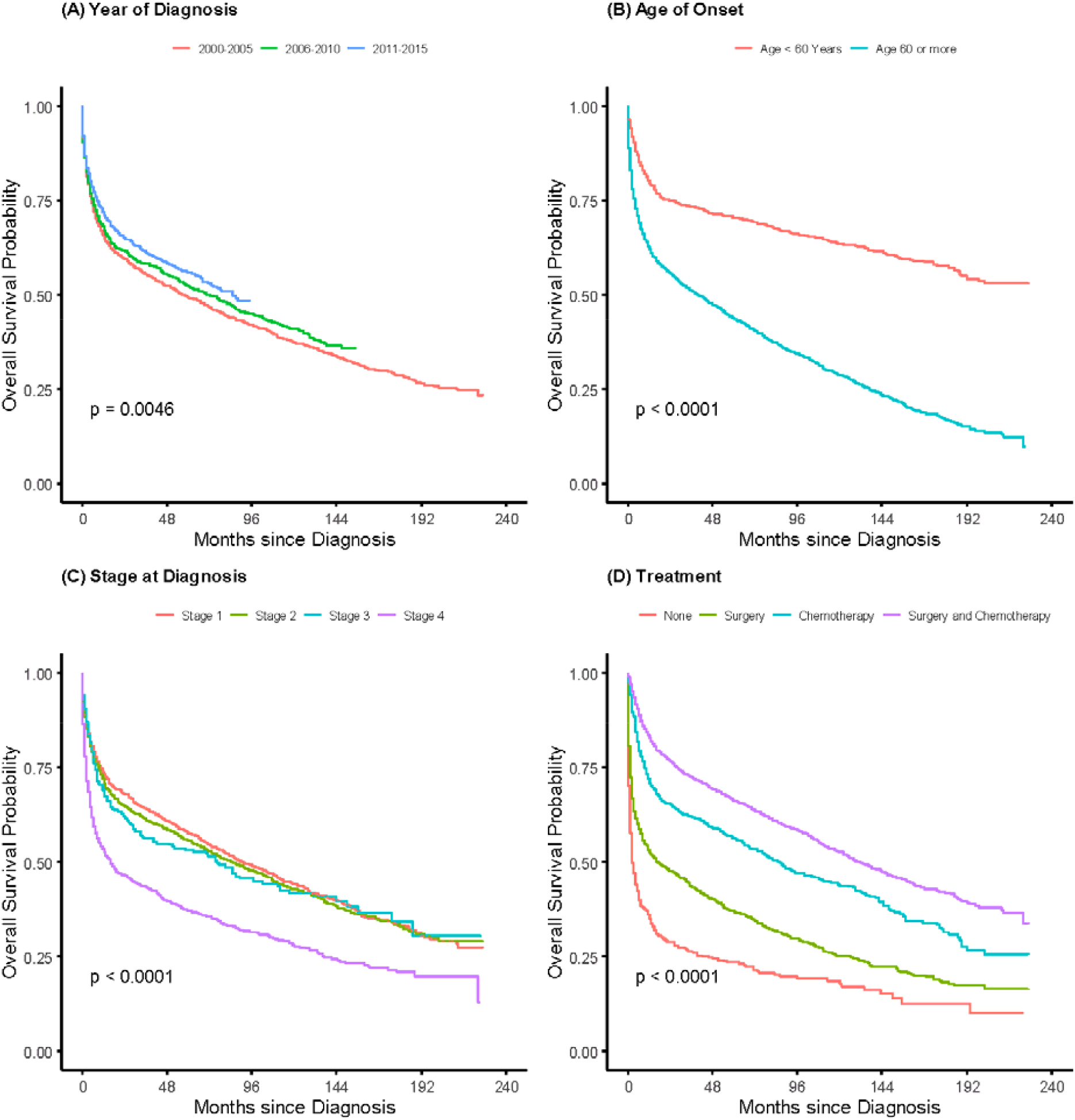
Kaplan Meier plots for Overall survival probability for Primary intestinal DLBCL for effects: (A) Year of Diagnosis, (B) Age, (d) Stage of disease and (D) treatment.

**Figure 5.**
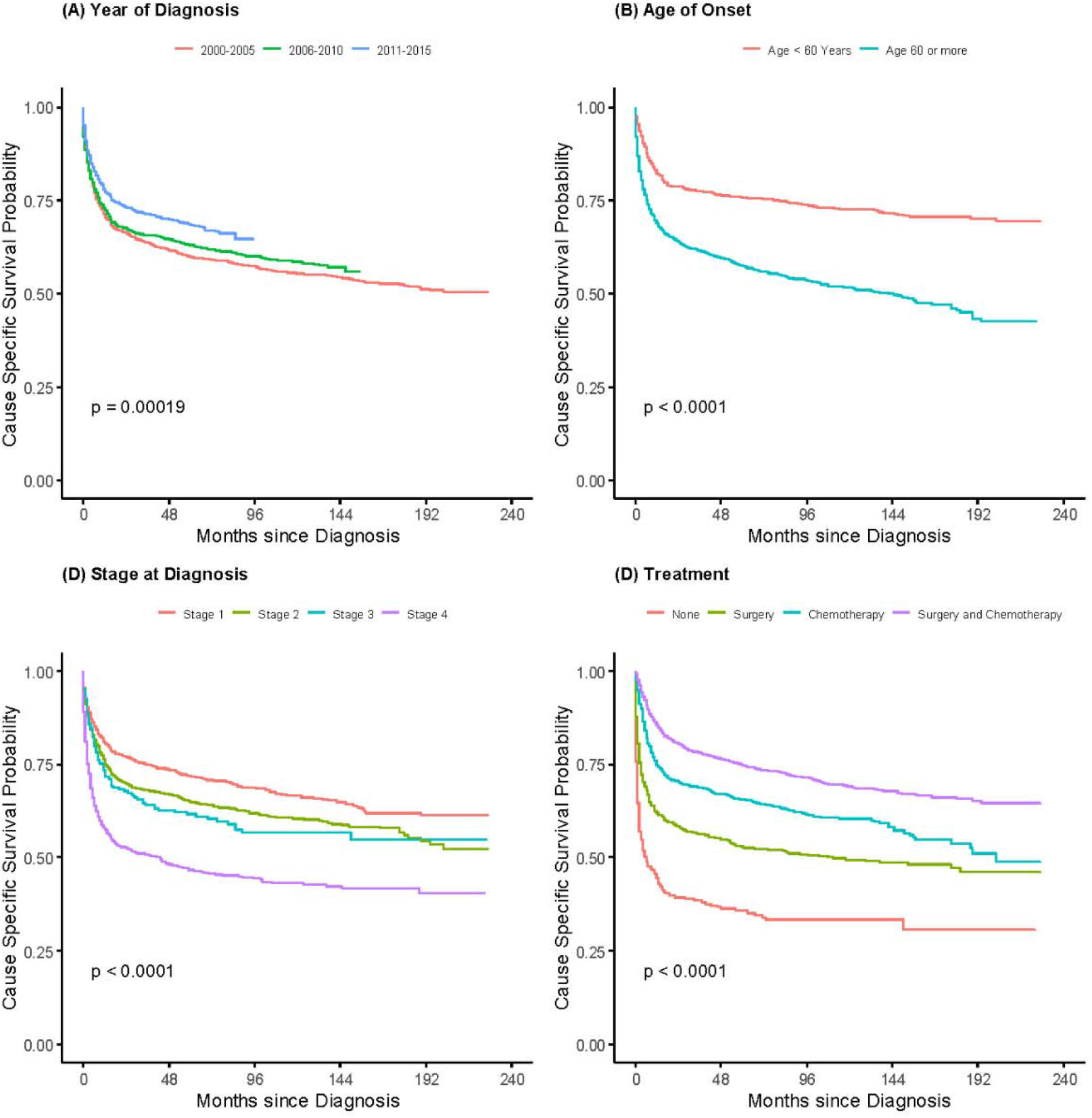
Kaplan Meier plot showing Cause Specific survival probability for PI-DLBCL for effects: (A) Year of Diagnosis and (B) Age, (d) Stage of disease and (D) treatment.

**Table 6.**
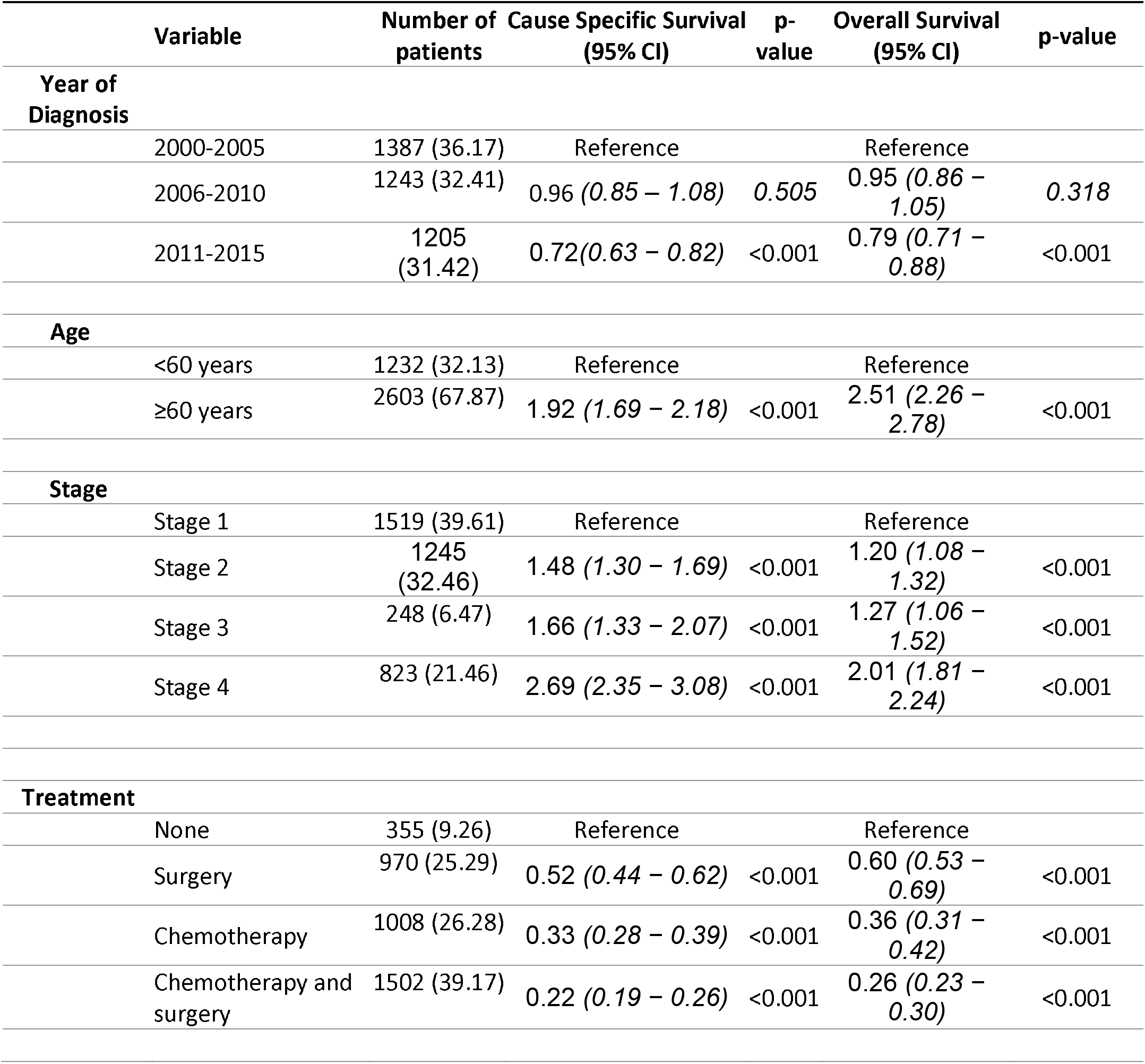
Multivariate Cox-proportional hazard analysis of the survival factors for cause specific and overall survival for PI-DLBCL patients.

## 4. Discussion

Although PI-NHL is a rare cancer, it accounts for a considerable portion of extra nodal NHL lymphomas. The SEER database of the National Cancer Institute which enlists data for cancer patients representing 27.8% percent of the US population is an excellent resource for doing a large population-based study for this rare entity. To the best of our knowledge, our study with more than 9000 PINHL patients is the largest analysis of clinical characteristics and outcomes among patients with PINHL.

Although the incidence of PI-NHL has remained relatively same over the years of analysis between 2000 to 2015, CSS and OS have improved significantly. This may be attributed to the improvement in diagnosis and treatment of lymphoma in general. In our study, 65.77% of patients were aged ≥ 60 years with the median age of onset being 66.5 years (95% CI 55 – 77). The higher prevalence of the condition in patients >60 years c PINHL was more common among males in congruence with previous data. This is the only study which has reported population-based age-adjusted incidence rate with incidence rate ratio close to twice for male compared to female, 0.62 times for non-Hispanic black and 0.92 times for Hispanic patients compared to non-Hispanic white population. In the previous studies, the number of reported cases were almost twice in the males for the eastern Asian population to 1.5 times in German and North American populations, which were seen in our patient group as well (9,11,12).

On multivariate analysis, age, gender, stage of disease, histology, and type of treatment were found to be the factors affecting overall survival. Age of onset and stage is currently part of the IPI scoring for NHL. In our study, patients older than 60 years were at twice the risk of cause specific mortality, whereas overall survival was even more strongly dependent on age with 2.68 times worse outcome in this group. In a study done on the Medicare beneficiaries with newly diagnosed NHLs(13), it was seen that NHL-specific mortality increases over the age for all types of NHLs and is more influenced by the comorbidities for younger individuals than that of older patients. The presence of comorbidities influences mortality from other conditions. In terms of gender, male patients have higher risk of both cause specific and overall mortality for unknown reasons.

We did not observe any significant difference in outcome between the small and large intestine primary sites. DLBCL is the most common subtype of PI-NHL, followed by follicular lymphoma and MALToma. DLBCL and BL had similar incidence in small and large intestine, whereas MALTOMA and Mantle cell lymphoma was more common in the large intestine (Table 2). Similar findings were seen in previous studies comprising data from multiple centers (12,14,15). While the median OS of primary intestinal T-cell lymphoma was 6 months, and PI-DLBCL 71 months, it was not reached in follicular lymphoma and was 165 months in MALToma. CSS was 8 months in T-cell lymphoma indicating the disease as cause of death in majority of patients, while the median CSS was not reached in other common histologies.

Treatment of PI-NHL varies based on histologic subtypes, clinical presentation, stage, and site. Indolent NHL like Follicular lymphoma and Mantle cell lymphoma can be managed by observation alone, as seen in 19% of our patient population(16). Chemotherapy is the primary modality of treatment for all types of NHL and holds true for intestinal NHL. In our patient population, more than half of all patients and more than half of patients who underwent surgery received chemotherapy, as seen in a recent study published on small intestinal lymphoma utilizing the National cancer database(17). This could indicate that many PI-NHL presents with intestinal mass and symptoms of obstruction, which necessitates the use of surgical intervention for immediate management. In a report published from China, around 48% of patients with PI-NHL required surgical intervention(12). In a recent report published from the SEER database on duodenal NHL, there has been a decline in the proportion of patients requiring surgery for duodenal NHL. In our study, which incorporated small and large intestine NHL, with around 60% of patients required surgical intervention.

Patients who underwent multimodality treatment with surgery and chemotherapy had better outcomes, followed by patients who received chemotherapy. Patients who underwent surgical management alone, although had a statistically significant improved CSS compared to patients who did not receive any treatment, there was no OS benefit noted. In a retrospective study which showed similar outcomes, it was observed that patients receiving surgery and chemotherapy were younger, male, and patients with fewer comorbidities(18–20). Another important reason that was highlighted for better survival in this group compared to those receiving chemotherapy alone is the low local relapse rate in intestinal B-cell lymphoma (21).

Similar findings of were also noted on our separate analysis of PI-DLBCL. Effects of treatment was more evident here with patients receiving chemotherapy with or without surgery having better CSS and OS compared to those who received surgery alone. Patient older than 60 years had worse outcomes compared to younger patients. Since the benefit of chemotherapy is evident in these patients, comorbidities, age, and surgical complications could have precluded them from receiving systemic chemotherapy contributing to the inferior survival. The overall outcomes have improved in recent years (2011-2015) and could be from introduction of rituximab, use of tolerable chemotherapy regimens and better management of chemotherapy related complications (22).

Considering the rare occurrence of this subtype of intestinal cancer, large population-based database like SEER database has been utilized to provide conclusive epidemiological data. However, our study has certain limitations. This is a retrospective study with registry data lacking detailed analysis of clinical characteristics, biomarkers including cytogenetics data, and complete treatment. Similarly cause of death data which is collected from death certificate in ordered to identify single, disease specific cause of death has some limitations with risk of misattributions(23). It has been also seen that SEER database has limited utility with low sensitivity in identifying cancer treatment which varied by cancer site, stage, and patient characteristics(24). Although positive predictive value was >85% for majority of treatment and cancer type which has improved over the time in recent studies(25).

## 5. Conclusions

This is the largest study reported on PINHL and summarizes the epidemiology of the primary intestinal Hodgkin’s lymphoma and factors that influence survival. Our data suggest that the evaluation of PI-NHL based on epidemiology, site and histological subtype remains very informative and very much needed. In our study we found that PI-NHL is mainly a disease of old age with higher incidence in non-Hispanic White, and male patient. Male gender, late-stage tumors and certain histologic types including T-cell lymphoma, BL and DLBCL are associated with poor outcomes. We also concluded that the outcome of combination of surgery and chemotherapy is not significantly different from chemotherapy alone but has better outcome than surgery or observation/no treatment. Further studies conducted by multicenter collaboration encompassing important details along with biomarkers will add lots of value to this subtype on NHL and rare type of intestinal cancer. A similar study with detailed data collection from SEER-Medicare database would also help in bridging some of the gap as well.

## Data Availability

All data produced in the present study are available upon reasonable request to the authors.

https://seer.cancer.gov/data-software/

## Acknowledgements

Not Applicable

## Author Contributions

Conceptualization, V.S, V.G, A.J, and T.M; methodology, V.S, D.G., V.G, A.J, D.D, H.E. and T.M.; data curation, V.S, V.G, and A.J; formal analysis, V.S, V.G, A.J, and D.G; original draft preparation V.S, D.G., V.G, A.J., D.D., H.E, and T.M; writing—review and editing, V.S, D.G., V.G, A.J., D.D., H.E, and T.M.; All authors have contributed equally, read and agreed to the published version of the manuscript.

## Funding

This research did not receive any external funding. APC will be funded by institutional fund.

## Financial Disclosure

There was no specific funding source to be mentioned

## Conflict of Interest

The authors declare that they have no competing interest.

## Ethical approval statement

Deidentified patient data were collected from SEER Database under data user agreement with NCI. Ethical review and approval were waived for this study, due to the de-identified information of the patients included in the public Surveillance, Epidemiology, and End Results database.

## Data availability statement

All data were extracted from and available on the Surveillance, Epidemiology, and End Results Program (https://seer.cancer.gov/, accessed on 10 September 2021

## Notes

### Competing Interest Statement

The authors have declared no competing interest.

### Funding Statement

This study did not receive any funding

### Author Declarations

Deidentified patient data were collected from Surveillance, Epidemiology, and End results Database under data user agreement with National Cancer Institute. Ethical review and approval were waived for this study, due to the de-identified information of the patients included in the public Surveillance, Epidemiology, and End Results database.

