## Supplement 1 for "Epidemiology and Determinants of Survival for Primary Intestinal Non-Hodgkin’s Lymphoma – A Population Based Study"

**Short title: Epidemiology of Primary intestinal non-Hodgkin’s lymphoma**

Vinit Singh^a^, Dhairya Gor^b^, Varsha Gupta^c,g^, Aasems Jacob^d^, Doantrang Du^a^, Hussam Eltoukhy^e^, Trishal Meghal^f^

**Supplement 1:**

**A1**: Selection Algorithm used for calculating age-adjusted incidence of Primary intestinal Extranodal Non-Hodgkin’s Lymphoma in Rate Session of SEER Stat:

Database: Incidence – SEER Research Data, 18 Registries, Nov 2020 Sub (2000-2018)

Statistics: Age-Adjusted Incidence Rate

Standard population: 2000 US Standard population (single ages to 84 -Census P25-1130)

Age Variable: Age recode with single ages and 85+

Selection Parameters:

{Behavior code ICD-O-3} = 'Malignant'

AND {Age recode with single ages and 85+} = '00 years', '01 years', '02 years', '03 years', '04 years', '05 years', '06 years', '07 years', '08 years', '09 years', '10 years', '11 years', '12 years', '13 years', '14 years', '15 years', '16 years', '17 years', '18 years', '19 years', '20 years', '21 years', '22 years', '23 years', '24 years', '25 years', '26 years', '27 years', '28 years', '29 years', '30 years', '31 years', '32 years', '33 years', '34 years', '35 years', '36 years', '37 years', '38 years', '39 years', '40 years', '41 years', '42 years', '43 years', '44 years', '45 years', '46 years', '47 years', '48 years', '49 years', '50 years', '51 years', '52 years', '53 years', '54 years', '55 years', '56 years', '57 years', '58 years', '59 years', '60 years', '61 years', '62 years', '63 years', '64 years', '65 years', '66 years', '67 years', '68 years', '69 years', '70 years', '71 years', '72 years', '73 years', '74 years', '75 years', '76 years', '77 years', '78 years', '79 years', '80 years', '81 years', '82 years', '83 years', '84 years', '85+ years'

{Age at Diagnosis.Age recode with single ages and 85+} = '18 years','19 years','20 years','21 years','22 years','23 years','24 years','25 years','26 years','27 years','28 years','29 years','30 years','31 years','32 years','33 years','34 years','35 years','36 years','37 years','38 years','39 years','40 years','41 years','42 years','43 years','44 years','45 years','46 years','47 years','48 years','49 years','50 years','51 years','52 years','53 years','54 years','55 years','56 years','57 years','58 years','59 years','60 years','61 years','62 years','63 years','64 years','65 years','66 years','67 years','68 years','69 years','70 years','71 years','72 years','73 years','74 years','75 years','76 years','77 years','78 years','79 years','80 years','81 years','82 years','83 years','84 years','85+ years'

{Race, Sex, Year Dx.Year of diagnosis} = '2000','2001','2002','2003','2004','2005','2006','2007','2008','2009','2010','2011','2012','2013','2014','2015'

{Site and Morphology.Site recode ICD-O-3/WHO 2008} = '	NHL - Extranodal'

AND {Site and Morphology.Primary Site - labeled} = 'C17.0-Duodenum','C17.1-Jejunum','C17.2-Ileum','C17.3-Meckels diverticulum','C17.8-Overlapping lesion of small intestine','C17.9-Small intestine, NOS','C18.0-Cecum','C18.1-Appendix','C18.2-Ascending colon','C18.3-Hepatic flexure of colon','C18.4-Transverse colon','C18.5-Splenic flexure of colon','C18.6-Descending colon','C18.7-Sigmoid colon','C18.8-Overlapping lesion of colon','C18.9-Colon, NOS','C19.9-Rectosigmoid junction','C20.9-Rectum, NOS','C21.0-Anus, NOS','C21.1-Anal canal','C21.2-Cloacogenic zone','C21.8-Overlapping lesion of rectum, anus, and anal canal'

**A2:** Selection Algorithm used for identifying Primary intestinal Extranodal Non-Hodgkin’s Lymphoma in Case listing Session of SEER Stat:

{Behavior code ICD-O-3} = 'Malignant'

AND {Age recode with <1 year olds} = '00 years', '01-04 years', '05-09 years', '10-14 years', '15-19 years', '20-24 years', '25-29 years', '30-34 years', '35-39 years', '40-44 years', '45-49 years', '50-54 years', '55-59 years', '60-64 years', '65-69 years', '70-74 years', '75-79 years', '80-84 years', '85+ years'

{Race, Sex, Year Dx.Year of diagnosis} = '2000','2001','2002','2003','2004','2005','2006','2007','2008','2009','2010','2011','2012','2013','2014','2015'

AND {Race and Age (case data only).Age recode with single ages and 85+} = '18 years','19 years','20 years','21 years','22 years','23 years','24 years','25 years','26 years','27 years','28 years','29 years','30 years','31 years','32 years','33 years','34 years','35 years','36 years','37 years','38 years','39 years','40 years','41 years','42 years','43 years','44 years','45 years','46 years','47 years','48 years','49 years','50 years','51 years','52 years','53 years','54 years','55 years','56 years','57 years','58 years','59 years','60 years','61 years','62 years','63 years','64 years','65 years','66 years','67 years','68 years','69 years','70 years','71 years','72 years','73 years','74 years','75 years','76 years','77 years','78 years','79 years','80 years','81 years','82 years','83 years','84 years','85+ years'

AND {Site and Morphology.Primary Site - labeled} = 'C17.0-Duodenum','C17.1-Jejunum','C17.2-Ileum','C17.3-Meckels diverticulum','C17.8-Overlapping lesion of small intestine','C17.9-Small intestine, NOS','C18.0-Cecum','C18.1-Appendix','C18.2-Ascending colon','C18.3-Hepatic flexure of colon','C18.4-Transverse colon','C18.5-Splenic flexure of colon','C18.6-Descending colon','C18.7-Sigmoid colon','C18.8-Overlapping lesion of colon','C18.9-Colon, NOS','C19.9-Rectosigmoid junction','C20.9-Rectum, NOS','C21.0-Anus, NOS','C21.1-Anal canal','C21.2-Cloacogenic zone','C21.8-Overlapping lesion of rectum, anus, and anal canal'

AND {Site and Morphology.Site recode ICD-O-3/WHO 2008} = '	NHL - Extranodal'

| Primary Site | Total Number |
| --- | --- |
| C17.0-Duodenum | 1301 |
| C17.1-Jejunum | 698 |
| C17.2-Ileum | 1053 |
| C17.3-Meckels diverticulum | 0 |
| C17.8-Overlapping lesion of small intestine | 98 |
| C17.9-Small intestine, NOS | 1828 |
| C18.0-Cecum | 1202 |
| C18.1-Appendix | 125 |
| C18.2-Ascending colon | 529 |
| C18.3-Hepatic flexure of colon | 77 |
| C18.4-Transverse colon | 219 |
| C18.5-Splenic flexure of colon | 44 |
| C18.6-Descending colon | 146 |
| C18.7-Sigmoid colon | 425 |
| C18.8-Overlapping lesion of colon | 117 |
| C18.9-Colon, NOS | 520 |
| C19.9-Rectosigmoid junction | 107 |
| C20.9-Rectum, NOS | 592 |
| C21.0-Anus, NOS | 34 |
| C21.1-Anal canal | 11 |
| C21.8-Overlapping lesion of rectum, anus, and anal canal | 17 |
| **Grand Total** | **9143** |

**ST1:** WHO-ICD-O Primary Site codes and distribution of PINHL Across the intestinal Tract

| **Year of Diagnosis** | **Age-Adjusted Incidence** | **Patient Count** | **Population** |
| --- | --- | --- | --- |
| 2000 | 0.8783 | 479 | 58040694 |
| 2001 | 0.9064 | 503 | 58817050 |
| 2002 | 0.8824 | 495 | 59501586 |
| 2003 | 0.8793 | 504 | 60151971 |
| 2004 | 1.0223 | 598 | 60813148 |
| 2005 | 0.9024 | 531 | 61314351 |
| 2006 | 0.8727 | 525 | 61900756 |
| 2007 | 0.9852 | 599 | 62575462 |
| 2008 | 1.0037 | 625 | 63371313 |
| 2009 | 0.8978 | 576 | 64169315 |
| 2010 | 0.9848 | 638 | 64917598 |
| 2011 | 0.9365 | 620 | 65640236 |
| 2012 | 0.8804 | 605 | 66346324 |
| 2013 | 0.8905 | 619 | 66988756 |
| 2014 | 0.8575 | 608 | 67632233 |
| 2015 | 0.8465 | 618 | 68266723 |
| **2000-2015** | **0.9145** | **9143** | **1010447516** |

**ST2:** Annual age-adjusted incidence rate from year 2000-2015; adjusted for 2000 US Standard population (Standard population for study population: 203,852,188)

|  | **Variable** | **Univariate cause Specific Survival HR (95% CI)** | **p-value** | **Univariate overall Survival**  **HR (95% CI)** | **p-value** |
| --- | --- | --- | --- | --- | --- |
| **Age**** |  |  |  |  |  |
|  | <60 years | Reference |  | Reference |  |
|  | ≥60 years | 2.07 (1.89 – 2.27) | <0.001 | 2.81 (2.61 – 3.03) | <0.001 |
| **Gender**** |  |  |  |  |  |
|  | Female | Reference |  | Reference |  |
|  | Male | 1.21 (1.11 – 1.31) | <0.001 | 1.14 (1.07 – 1.21) | <0.001 |
| **Ethnicity^¥^** |  |  |  |  |  |
|  | Non-Hispanic white | Reference |  | Reference |  |
|  | Non-Hispanic black | 1.21 (1.04 – 1.41) | 0.01 | 1.18 (1.04 – 1.32) | 0.007 |
|  | Hispanic | 1.04 (0.92 – 1.18) | 0.52 | 0.90 (0.81 – 0.99) | 0.039 |
|  | Others | 0.98 (0.85 – 1.12) | 0.74 | 0.90 (0.81 – 1.00) | 0.048 |
| **Staging**** |  |  |  |  |  |
|  | Stage 1 | Reference |  | Reference |  |
|  | Stage 2 | 1.64 (1.48 – 1.80) | <0.001 | 1.25 (1.16 – 1.34) | <0.001 |
|  | Stage 3 | 1.92 (1.63 – 2.26) | <0.001 | 1.38 (1.21 – 1.58) | <0.001 |
|  | Stage 4 | 2.65 (2.40 – 2.91) | <0.001 | 1.83 (1.70 – 1.98) | <0.001 |
| **Site^¥^** |  |  |  |  |  |
|  | Small Intestine | Reference |  | Reference |  |
|  | Large Intestine | 0.95 (0.88 – 1.03) | 0.23 | 1.04 (0.98 – 1.11) | 0.1 |
| **Histology**** |  |  |  |  |  |
|  | DLBCL | Reference |  | Reference |  |
|  | Follicular Lymphoma | 0.22 (0.19 – 0.26) | <0.001 | 0.35 (0.31 – 0.38) | <0.001 |
|  | MALTOMA | 0.25 (0.22 – 0.30) | <0.001 | 0.47 (0.43 – 0.53) | <0.001 |
|  | Mantle Cell lymphoma | 0.88 (0.75 – 1.03) | 0.11 | 0.83 (0.73 – 0.94) | 0.004 |
|  | Other B-cell NHL | 0.87 (0.70 – 1.10) | 0.25 | 0.99 (0.83 – 1.17) | 0.870 |
|  | NHL, NOS | 0.90 (0.77 – 1.04) | 0.15 | 0.98 (0.88 – 1.10) | 0.765 |
|  | Burkitt Lymphoma | 1.00 (0.83 – 1.19) | 0.96 | 0.75 (0.65 – 0.89) | <0.001 |
|  | T−Cell NHL | 2.96 (2.61 – 3.37) | <0.001 | 2.42 (2.15 – 2.72) | <0.001 |
| **Treatment**** |  |  |  |  |  |
|  | None | Reference |  | Reference |  |
|  | Surgery | 1.21 (1.08 – 1.37) | 0.001 | 1.31 (1.20 – 1.43) |  |
|  | Chemotherapy | 1.13 (1.00 – 1.27) | 0.54 | 1.00 (0.91 – 1.10) | 1.00 |
|  | Chemotherapy and surgery | 0.98 (0.87 – 1.10) | 0.70 | 0.87 (0.79 - .95) | 0.002 |
| **Year of Diagnosis**** |  |  |  |  |  |
|  | 2000 – 2005 | Reference |  | Reference |  |
|  | 2006 - 2010 | 0.81 (0.74 – 0.89) | <0.001 | 0.84 (0.78 – 0.90) | <0.001 |
|  | 2011 - 2015 | 0.69 (0.63 – 0.77) | <0.001 | 0.76 (0.70 – 0.82) | <0.001 |

**ST3:** Univariate analysis of the survival factors for cause specific and overall survival for all PINHL patients. ¥ Cause specific cox proportional hazard model was not significant for site (p-value – 0.2) and ethnicity (p-value – 0.1). Hence, Not included for multivariate analysis. **Cox PH model p-value – <0.001.

| **Histology** | **Stage** | | | | **Treatment** | | | |
| --- | --- | --- | --- | --- | --- | --- | --- | --- |
|  | **Stage 1** | **Stage 2** | **Stage 3** | **Stage 4** | **None** | **Surgery** | **Chemotherapy** | **Both** |
| **DLBCL** | 1,519 (39.61) | 1,245 (32.46) | 238 (6.47) | 823 (21.46) | 355 (9.26) | 970 (25.29) | 1,008 (26.28) | 1,502 (39.17) |
| **Follicular Lymphoma** | 870 (55.66) | 414 (26.49) | 66 (4.22) | 213 (13.63) | 503 (32.18) | 495 (31.67) | 275 (17.59) | 290 (18.55) |
| **MALTOMA** | 812 (70.00) | 189 (16.29) | 34 (2.93) | 125 (10.78) | 368 (31.72) | 516 (44.48) | 155 (13.36) | 121 (10.43) |
| **Mantle Cell lymphoma** | 138 (30.87) | 65 (14.54) | 37 (8.28) | 207 (46.31) | 89 (19.91) | 60 (13.42) | 197 (44.07) | 101 (22.60) |
| **Other B-cell NHL** | 100 (43.10) | 58 (25.00) | 14 (6.03) | 60 (25.86) | 59 (25.43) | 80 (34.48) | 51 (21.98) | 42 (18.10) |
| **NHL, NOS** | 327 (54.96) | 132 (22.18) | 20 (3.36) | 116 (19.50) | 168 (28.24) | 158 (26.55) | 166 (27.90) | 103 (17.31) |
| **Burkitt Lymphoma** | 100 (29.07) | 109 (31.69) | 11 (3.20) | 124 (36.05) | 18 (5.23) | 41 (11.92) | 98 (28.49) | 187 (54.36) |
| **T−Cell NHL** | 147 (37.50) | 117 (29.85) | 26 (6.63) | 102 (26.02) | 38 (9.69) | 126 (32.14) | 68 (17.35) | 160 (40.82) |
| **Total** | 4,013 (46.84) | 2,329 (27.18) | 456 (5.32) | 1,770 (20.66) | 1,598 (18.65) | 2,446 (28.55) | 2,018 (23.55) | 2,506 (29.25) |

**ST4**: Stage and Treatment distribution of each histological subtype of PINHL
